# Probabilistic approaches for classifying highly variable anti-SARS-CoV-2 antibody responses

**DOI:** 10.1101/2020.07.17.20155937

**Authors:** Xaquin C Dopico, Leo Hanke, Daniel J. Sheward, Sandra Muschiol, Soo Aleman, Nastasiya F. Grinberg, Monika Adori, Murray Christian, Laura Perez Vidakovics, Changil Kim, Sharesta Khoenkhoen, Pradeepa Pushparaj, Ainhoa Moliner Morro, Marco Mandolesi, Marcus Ahl, Mattias Forsell, Jonathan Coquet, Martin Corcoran, Joanna Rorbach, Joakim Dillner, Gordana Bogdanovic, Gerald M. McInerney, Tobias Allander, Ben Murrell, Chris Wallace, Jan Albert, Gunilla B. Karlsson Hedestam

**Author notes:** Equal contribution (Leo Hanke, Daniel J. Sheward, Sandra Muschiol, Soo Aleman).

## Abstract

Antibody responses vary widely between individuals^1^, complicating the correct classification of low-titer measurements using conventional assay cut-offs. We found all participants in a clinically diverse cohort of SARS-CoV-2 PCR+ individuals (*n*=105) – and *n=*33 PCR+ hospital staff – to have detectable IgG specific for pre-fusion-stabilized spike (S) glycoprotein trimers, while 98% of persons had IgG specific for the receptor-binding domain (RBD). However, anti-viral IgG levels differed by several orders of magnitude between individuals and were associated with disease severity, with critically ill patients displaying the highest anti-viral antibody titers and strongest *in vitro* neutralizing responses. Parallel analysis of random healthy blood donors and pregnant women (*n=*1,000) of unknown serostatus, further demonstrated highly variable IgG titers amongst seroconverters, although these were generally lower than in hospitalized patients and included several measurements that scored between the classical 3 and 6SD assay cut-offs. Since the correct classification of seropositivity is critical for individual- and population-level metrics, we compared different probabilistic algorithms for their ability to assign likelihood of past infection. To do this, we used tandem anti-S and -RBD IgG responses from our PCR+ individuals (*n=*138) and a large cohort of historical negative controls (*n=*595) as training data, and generated an equal-weighted learner from the output of support vector machines and linear discriminant analysis. Applied to test samples, this approach provided a more quantitative way to interpret anti-viral titers over a large continuum, scrutinizing measurements overlapping the negative control background more closely and offering a probability-based diagnosis with potential clinical utility. Especially as most SARS-CoV-2 infections result in asymptomatic or mild disease, these platform-independent approaches improve individual and epidemiological estimates of seropositivity, critical for effective management of the pandemic and monitoring the response to vaccination.

## Introduction

The characterization of nascent SARS-CoV-2-specific antibody responses is critical to our understanding of the infection at individual and population levels. Although several groups have carried out elegant work in this regard^2–5^, consensus on several key issues remains outstanding, such as: whether all infected persons develop an antibody response to the virus; what the duration of these responses is following peak levels; and what titers provide protective immunity against re-infection^6–8^.

Antibody responses to the SARS-CoV-2 spike glycoprotein (S) are particularly relevant, as S-directed antibody specificities mediate virus neutralizing activity and new S variants (such as B.1.1.7) have emerged. Indeed, the vast majority of COVID-19 vaccines are based on S surface antigens, as the goal is to induce neutralizing antibodies that block viral entry into ACE2-positive target cells^9,10^. Furthermore, because serological studies play such a central role in immunosurveillance, there is a pressing need for robust assays and quantitative statistical tools to examine antibody titers of varying levels after vaccination and natural infection in different target groups.

To meet these needs, we developed highly sensitive and specific IgM, IgG and IgA ELISA assays based on mammalian cell-expressed pre-fusion-stabilized soluble trimers of the SARS-CoV-2 spike (S) glycoprotein and the receptor-binding domain (RBD), and used them in tandem to survey serum samples from large cohort of individuals PCR+ for SARS-CoV-2. To validate our assays, we repeatedly analyzed a large set of serum samples from historical blood donors as negative controls (*n=*595) - critical for determining the assay background.

As we show, and as has been reported by others^4,7,11^, the magnitude of response varied greatly between seropositive individuals and was associated with disease severity. Those with most pronounced symptoms had the highest anti-viral antibody titers, while those with asymptomatic or mild disease (including otherwise healthy blood donors and pregnant women) exhibited a range of antibody levels, with many measurements in close approximation to the negative control background, complicating their correct classification. To improve upon the dichotomization of a continuous variable – which is common to many clinical tests but results in a loss of information^12,13^ – we used tandem anti-S and RBD IgG data from confirmed infections and negative controls to train different probabilistic algorithms to assign likelihood of past infection. Compared to strictly thresholding the assay at 3 or 6 standard deviations (SD) from the mean of negative control measurements, these more quantitative approaches modelled the probability a sample was positive, improving the identification of low titer values and paving the way for a greater utility to antibody test results.

## Results

Study samples are detailed in Fig. 1A and Table 1.

**Table 1.** Study samples.

|  |  |
| --- | --- |
| **SARS-CoV-2 PCR+ individuals<sup>§</sup>** | *n*=105 |
| Females | 44 (41.9%) |
| Males | 61 (58.1%) |
| Median age (years) | 53.0 (49-61) |
| Females | 51.5 (48-56.2) |
| Males | 55.0 (49-63) |
| Non-hospitalized (*n*=) | 53 |
| Females, males | 28, 25 |
| Hospitalized patients (*n*=) | 31 |
| Females, males | 12, 17 |
| Intensive care (ICU) patients (*n*=) | 21 |
| Females, males | 3, 17 |
| SARS-CoV2+ PCR (*n*=) | 105 |
| Sample collection dates | March-May 2020 |
| **SARS-CoV-2 PCR+ KI hospital staff \*** | *n*=33 |
| Sample collection dates | July 2020 |
| **Blood donors \*** | *n*=500 |
| Sample collection dates | Weeks 17-21 (March-May) 2020 |
| **Pregnant women \*** | *n*=500 |
| Sample collection dates | Weeks 17-21 (March-May) 2020 |
| **Historical blood donors \*** | *n*=595 |
| Sample collection dates | March-June 2019 |
| **ECV+ donors \*** | *n*=20 |
| Sample collection dates | July-December 2019 |
| <sup>§</sup>Under the care of Karolinska University Hospital |  |
| *No additional metadata available for any samples |  |

**Figure 1:**
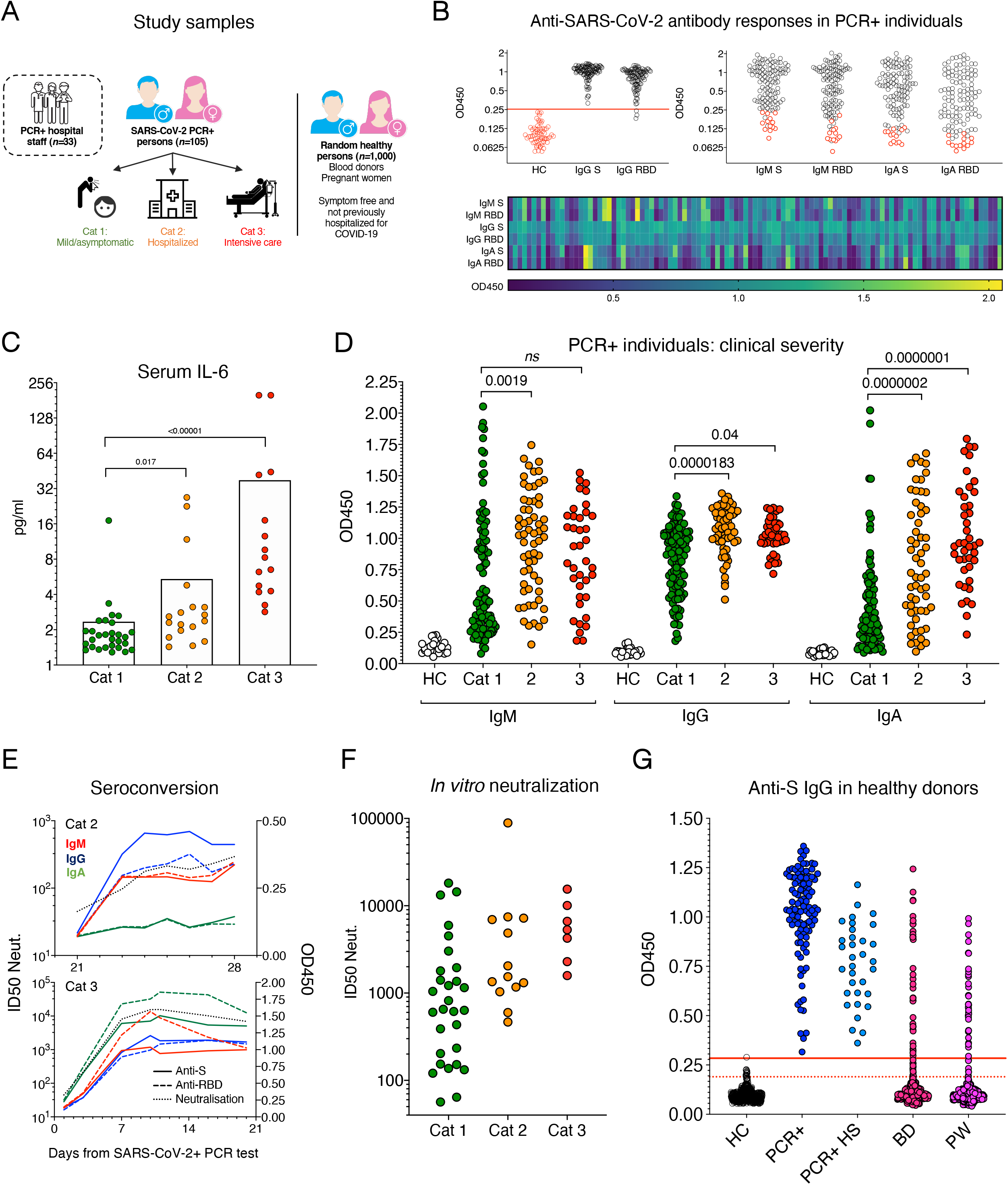
Anti-SARS-CoV-2 Ab phenotypes in COVID-19 patients, PCR+ individuals, blood donors and pregnant women. **(A)** Study samples. **(B)** Raw optical density (450nm) for anti-S and -RBD IgG (first graph) IgM and IgA responses (second graph) in SARS-CoV-2 PCR+ individuals (*n*=105). A 6SD cut-off based on *n=*595 control (HC) values is shown for the IgG assay and a small number of HC samples are shown (red) for the IgM and IgA assays. Individual responses for the three isotypes are shown by a heatmap. **(C)** Circulating IL-6 levels in serum are associated with disease severity. **(D)** Anti-viral antibody levels are associated with disease severity, most pronounced for anti-viral IgA. Anti-S and RBD responses are graphed together. *P* values for S are shown. **(E)** Two discordant longitudinal profiles of seroconversion and neutralisation capacity are shown in hospitalized COVID-19 patients. **(F)** *In vitro* pseudotyped virus neutralization ID_50_ titers are associated with disease severity, with the highest titers observed in Cat 3 (ICU) patients. *n*=48 SARS-CoV-2 PCR+ individuals were analyzed in duplicate. **(G)** Comparison of anti-S IgG levels between PCR+ individuals (*n*=105), PCR+ hospital staff (PCR+ HS, *n=*33), blood donors (BD, *n=*500) and pregnant women (PW, *n=*500). 3 and 6 SD cut-offs are shown by red lines.

### Antibody test development

We developed ELISA protocols to profile IgM, IgG and IgA specific for a pre-fusion-stabilized spike (S) glycoprotein trimer^14^, the RBD, and the nucleocapsid (N). Trimer conformation was confirmed in each batch by cryo-EM^15^ and a representative subset of study samples was used for assay development (Fig. S1A). In contrast to other studies reporting significant cross-reactivity to S in the UK population^16^, we did not observe reproducible IgG reactivity to S or RBD across all 595 historical controls in the study, although two individuals who were PCR-positive for endemic coronaviruses (ECV+) in the last six months displayed reproducible IgM reactivity to both SARS-CoV-2 N and S, and two 2019 blood donors (from *n=*72 tested) had low anti-S IgM reactivity (Fig. S1B). Thus, further investigation is required to establish the contribution of potential cross-reactive memory SARS-CoV-2 responses^17^.

Responses to S and the RBD were highly correlated and our assay revealed a greater than 1,000-fold difference in anti-viral IgG titers between Ab-positive individuals when examining serially diluted sera (Fig. S1C and D). In SARS-CoV-2 PCR+ individuals, anti-viral IgG titers were comparable for S (EC_50_=3,064; 95% CI [1,197 - 3,626]) and N (EC_50_=2,945; 95% CI [543 - 3,936]) and lower for RBD [EC_50_=1,751; 95% CI 966 - 1,595]. Notably, a subset (*ca*. 10%) of the SARS-CoV-2-confirmed individuals did not have detectable IgG responses against the SARS-CoV-2 nucleocapsid protein (N) (Fig. S1C), as previously reported^18^. Therefore, we did not explore responses to N further. These results highlight that the choice of antigen is critical for seropositivity estimates.

### Elevated anti-viral Ab titers and neutralizing responses are associated with increased disease severity

When screening samples from SARS-CoV-2 PCR+ individuals from whom clinical information was available (*n=*105), we detected potent IgG responses against S in 100% of participants, and against RBD in 97% of persons (Fig. 1B), supporting that natural infection engenders a robust B cell response in the majority of cases, as reported^8^. IgM and IgA responses were generally weaker and more variable and also spread over a large range (Fig. 1B).

To examine this further, PCR+ individuals were grouped according to their clinical status: non-hospitalized (Cat. 1), hospitalized (Cat. 2) or admitted to the intensive care unit (Cat. 3). To validate our clinical classification, we measured serum IL-6 levels in a random subset of PCR+ individuals (*n=*64). IL-6 feeds Ab production^19–22^, and as has been reported^23^, was increased in samples from individuals with severe disease (Fig. 1C). Furthermore, multivariate analyses (accounting for the effects of age, sex and days from symptom onset/PCR test) revealed increased anti-viral IgM, IgG and IgA to be associated with disease severity, as has been reported^7^ (Fig. 1C and S1D-E, Table S1). Severe disease was most strongly associated with virus-specific IgA, suggestive of mucosal pathology. We did not observe an association between ICU or IL-6 status and IgM levels, supporting that levels of the cytokine and IgA mark a more severe clinical course of COVID-19 (Fig. S1D). Anti-RBD IgA responses were slightly lower in non-hospitalized and hospitalized females compared to males, and trended similarly for S (Fig. S1D and Table S1), consistent with females developing less severe disease^4^.

Across all PCR+ individuals (sampled up to two months from PCR test), anti-viral IgG levels were maintained, while IgM and IgA decreased, in agreement with their circulating t_1/2_ and viral clearance (Fig. S1D and Table S1). In longitudinal patient samples (sequential sampling of PCR+ individuals in the study) where we observed seroconversion, IgM, IgG and IgA peaked with similar kinetics when all three isotypes developed, although IgA was not always generated in non-hospitalized or hospitalized individuals (Fig 1E), supporting a more diverse antibody response in severe disease.

To extend these observations, we characterized the *in vitro* virus neutralizing antibody response in PCR+ patients. Using an established pseudotype virus neutralization assay^24^, we detected neutralizing antibodies in the serum of all SARS-CoV-2 PCR+ individuals screened (*n=*48) (Fig. 1F). Neutralizing responses were not seen in samples before seroconversion or negative controls (Fig. 1E and F). A large range of neutralizing ID_50_ titers was apparent, with binding and neutralization being highly correlated (Fig. S1D). In agreement with the binding data, the strongest neutralizing responses were observed in samples from patients in intensive care (g.mean ID_50_=5,058; 95% CI [2,422 - 10,564]) (Fig 1E).

In healthy blood donors and pregnant women (*n=*1,000 collected between weeks 17-21 2020 – the same time as the patient cohort), who did not have signs or symptoms of COVID-19 for two weeks prior to sampling, and had not been hospitalized for COVID-19, IgG titers varied greatly but were generally lower than hospitalized COVID-19 patients, and were comparable to titers in PCR+ hospital staff (*n=*33) who also had never been hospitalized following infection (Fig. 1G).

### Probabilistic analyses of positivity

As SARS-CoV-2 results in asymptomatic or mild disease in the majority of cases, and antibody titers decline following peak responses and viral clearance, the correct classification of low titer values is critical to individual and population-level estimates of antibody-positivity for COVID-19. Indeed, several healthy donor test samples screen in this study had optical densities between the 3 and 6 SD cut-offs for both or a single antigen (Fig. 1G), highlighting the problem of assigning case to *low responder* values.

To further our understanding of the assay boundary, we repeatedly analyzed a large number of historical (SARS-CoV-2-negative) controls (blood donors from the spring of 2019, *n*=595) alongside test samples throughout the study. We considered the spread of negative values critical, since the use of a small and unrepresentative set of controls can lead to an incorrectly set threshold, which can considerably skew the seropositivity estimate. This is illustrated by the random sub-sampling of non-overlapping groups of negative controls, resulting in a 40% difference in the positivity estimate (Fig. 2A). Worryingly, many clinically approved tests use a ratio between a known positive and negative serum calibrator to classify seropositivity^25^, although we show here that these are highly variable within the population.

**Figure 2:**
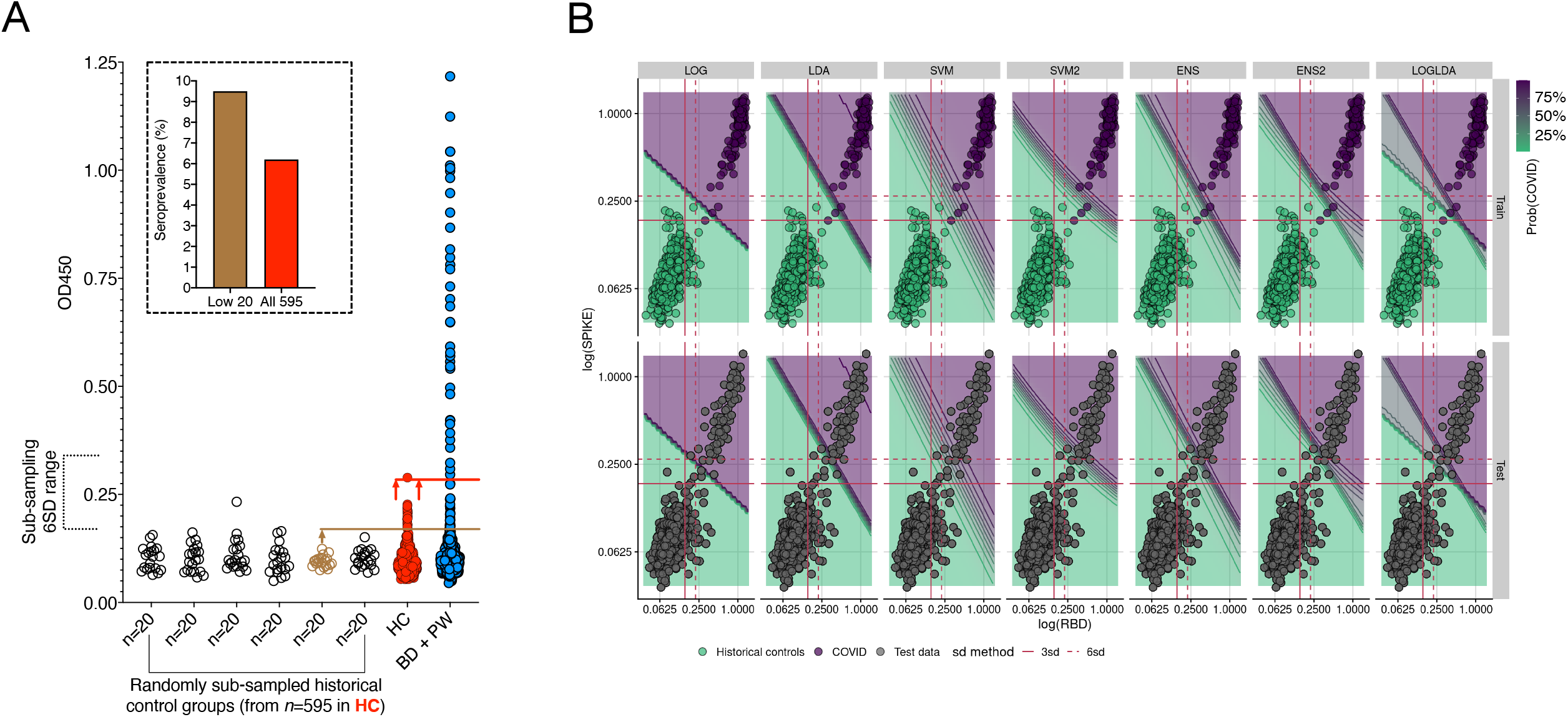
Probability-based seropositivity estimates in blood donors and pregnant women. **(A)** Random sub-sampling of non-overlapping negative controls illustrates how the range of negative control (C) values can influence a conventional test cut-off, here 6 SD from the mean of the respective C groups. In the test data, depending on the control values used to set the test threshold for positivity, SP estimates varied 40%. Blood donor and pregnant women sample values are used as an example. Anti-S IgG values are shown. **(B)** Comparison of probabilistic algorithms suited to ELISA measurements. Logistic regression (LOG), linear discriminant analysis (LDA), support vector machines (SVM) and quadratic SVM (SVM2). Learners were trained using anti-S and RBD IgG data from 595 negative control values and 138 SARS-CoV-2 PCR+ individual samples. Ensemble (*ENS*) learners were generated from the output of SVM-LDA, SVM2-LDA and LOG-LDA.

Therefore, to exploit individual titers generated against multiple antigens, we used anti-S and RBD data from PCR+ individuals and negative controls to train probabilistic algorithms to assign likelihood of past infection. To this end, we compared different probabilistic algorithms – logistic regression (LOG), linear discriminant analysis (LDA), linear support vector machines (SVM) and quadratic SVM (SVM2) – suited to ELISA data (Fig. 2B, Materials and Methods). Using ten-fold cross validation and training models on both proteins simultaneously (S and RBD), we found all methods worked well, with sensitivity >98% and specificity >99.6% (Fig. 2D). On these metrics, LDA gave the highest specificity. Logistic regression had similarly high specificity on some folds of the training data, but with higher sensitivity. However, we deliberately considered balanced and unbalanced folds (where case:control ratios varied between folds) and found LOG to show the least consistency across strategies, which reflects that the proportion of cases in a sample directly informs a logistic model’s estimated parameters. SVM methods had lower specificity than LDA in the training data, but higher sensitivity.

The standard methods, calling positives by a fixed number of SD above the mean of negative controls, displayed two extreme behaviors: 3-SD had the highest sensitivity (100%) while 6-SD had the highest specificity, and the lowest sensitivity (Fig. S2A), emphasizing that the number of SD above the mean is a key parameter, but one which is not learnt in any formal data-driven manner. Both SVM and LDA offer linear classification boundaries, but we can see that the probability transition from negative to positive cases is much sharper for LDA (Fig. 2B) – potentially resulting in false negatives when applied to the test data, but giving the model high specificity in the training data under cross-validation. SVM exhibits a softer probability transition around its classification boundary, offering a much more nuanced approach to the points lying in the mid-range of the two proteins. SVM2 creates a nonlinear boundary, but the cross validation suggested that this didn’t improve performance relative to linear SVM.

Given these results, we chose to create ensemble learners, which were unweighted averages of SVM (linear) or SVM2 (quadratic) and LDA (*ENS* and *ENS2*, respectively), as well as a LOG-LDA learner, to balance the benefits of each approach. The ensemble learners seemed to combine the benefits of their parent methods (Fig. S2A). Test data points in the lower right region of each plot are the hardest to classify due to the relative scarcity of observations in this region in the training dataset and *ENS* (SVM-LDA) showed the greatest uncertainty in these regions, appropriately. Given these results, we chose to use *ENS* (SVM-LDA), with an average sensitivity >99.1% and specificity >99.8%, to analyze test data. When applied to the serology data, the output of *ENS* is the probability of each sample being antibody-positive.

In healthy donor test data, the *ENS* learner estimated 7.8% (95% CI [4.8-12.5]) positivity in samples collected in week 21 of 2020 (Fig. S2B, Table S2). This is in contrast to the SD thresholding, which identified 12% and 10% positivity for S and RBD, respectively, at 3 SD, and 8% and 7.5, respectively, at 6 SD (Table S2). Therefore, apart from providing more accurate population-level estimates – critical to seroprevalence studies, where we have applied these and related tools in a large cohort^26^ – these methods have the potential to provide more nuanced information about titers to an individual after an antibody test. For example, test samples with a 30-60% chance of being antibody positive (Fig. S2B) can be targeted for further investigation or help inform vaccine boosting, as antibody titers decline over time from peak responses. Moreover, such tools are applicable to other clinical metrics where a continuous scale is dichotomized and code for implementation is freely available via our online repositories.

## Discussion

Benefitting from a robust antibody test developed alongside a diagnostic clinical laboratory responsible for monitoring sero-reactivity during the pandemic, we profiled SARS-CoV-2 antibody responses in three cohorts of clinical interest. COVID-19 patients receiving intensive care showed the highest anti-viral Ab titers, developing augmented serum IgA and IL-6 with worsening disease and more advance respiratory and/or gastrointestinal pathology. These results support the use of cytokine and isotype-level measures for patient management^23^.

Importantly, our neutralization data illustrated that nearly all SARS-CoV-2 PCR+ individuals developed neutralizing antibodies capable of preventing S-mediated cell entry, albeit at different titers. These data support that SARS-CoV-2 infection generates a functional B cell response in the majority of people^8^ and serve as a useful comparator to titers engendered by vaccination. Indeed, the first generation of mRNA vaccines have been reported to generate neutralizing titers comparable to samples from individuals with mild infection in our study^9,10^.

Outside of the severe disease setting, it is critical to accurately determine who and how many people have seroconverted for clinical and epidemiological reasons. However, this is complicated by low titer values, which in some cases – and increasingly with time since exposure or vaccination ^5,27^ – can overlap outlier values among negative control samples. Test samples with true low anti-viral titers fall into this range, highlighting the need to better understand the assay boundary. To improve upon strictly thresholding the assay, we developed probabilistic approaches for ELISA data that characterized the uncertainty in individual measures. These approaches provide more statistically sound measurements at the level of cohorts and the potential to communicate more nuanced information to individual patients – although the communication of probability needs to be approached with care to ensure what is described matches what an individual interprets. Furthermore, such approaches will aid the analysis of data from assay platforms measuring the responses to multiple antigens; longitudinal studies of the duration of immunity after SARS-CoV-2 spike-based vaccines and natural infection; and facilitate comparison of responses in different cohorts.

## Materials and methods

### Human samples and ethical declaration

Samples from PCR+ individuals and admitted COVID-19 patients (*n=*105) were collected by the attending clinicians and processed through the Departments of Medicine and Clinical Microbiology at the Karolinska University Hospital. Samples were used in accordance with approval by the Swedish Ethical Review Authority (registration no. 2020-02811). All personal identifiers were pseudo-anonymized, and all clinical feature data were blinded to the researchers carrying out experiments until data generation was complete. PCR testing for SARS-CoV-2 RNA was by nasopharyngeal swab or upper respiratory tract sampling at Karolinska University Hospital. As viral RNA levels were determined using different qPCR platforms (with the same reported sensitivity and specificity) between participants, we did not analyze these alongside other features. PCR+ individuals (*n=*105) were questioned about the date of symptom onset at their initial consultation and followed-up for serology during their care, up to 2 months post-diagnosis. Serum from SARS-CoV-2 PCR+ individuals was collected 6-61 days post-test, with the median time from symptom onset to PCR being 5 days. In addition, longitudinal samples from 10 of these patients were collected to monitor seroconversion and isotype persistence.

Hospital workers at Karolinska University Hospital were invited to test for the presence of SARS-CoV-2 RNA in throat swabs in April 2020 and virus-specific IgG in serum in July 2020. We screened 33 PCR+ individuals to provide additional training data for ML approaches. All participants provided written informed consent. The study was approved by the National Ethical Review Agency of Sweden (2020-01620) and the work was performed accordingly.

Anonymized samples from blood donors (*n=*100/week) and pregnant women (*n=*100/week) were randomly selected from their respective pools by the department of Clinical Microbiology, Karolinska University Hospital. No metadata, such as age or sex information were available for these samples in this study. Pregnant women were sampled as part of routine for infectious diseases screening during the first trimester of pregnancy. Blood donors (*n*=595) collected through the same channels a year previously were randomly selected for use as negative controls. Serum samples from individuals testing PCR+ for endemic coronaviruses, 229E, HKU1, NL63, OC43 (*n=*20, ECV+) in the prior 2-6 months, were used as additional negative controls. The use of study samples was approved by the Swedish Ethical Review Authority (registration no. 2020-01807). Stockholm County death and Swedish mortality data was sourced from the ECDC and the Swedish Public Health Agency, respectively. Study samples are defined in Table 1.

### Serum sample processing

Blood samples were collected by the attending clinical team and serum isolated by the department of Clinical Microbiology. Samples were anonymized, barcoded and stored at - 20°C until use. Serum samples were not heat-inactivated for ELISA protocols but were heat-inactivated at 56°C for 60 min for neutralization experiments.

### SARS-CoV-2 antigen generation

The plasmid for expression of the SARS-CoV-2 prefusion-stabilized spike ectodomain with a C-terminal T4 fibritin trimerization motif was obtained from^14^. The plasmid was used to transiently transfect FreeStyle 293F cells using FreeStyle MAX reagent (Thermo Fisher Scientific). The ectodomain was purified from filtered supernatant on Streptactin XT resin (IBA Lifesciences), followed by size-exclusion chromatography on a Superdex 200 in 5 mM Tris pH 8, 200 mM NaCl.

The RBD domain (RVQ – QFG) of SARS-CoV-2 was cloned upstream of a Sortase A recognition site (LPETG) and a 6xHIS tag, and expressed in 293F cells as described above. RBD-HIS was purified from filtered supernatant on His-Pur Ni-NTA resin (Thermo Fisher Scientific), followed by size-exclusion chromatography on a Superdex 200. The nucleocapsid was purchased from Sino Biological.

### Anti-SARS-CoV-2 ELISA

96-well ELISA plates (Nunc MaxiSorp) were coated with SARS-CoV-2 S trimers, RBD or nucleocapsid (100 μl of 1 ng/μl) in PBS overnight at 4°C. Plates were washed six times with PBS-Tween-20 (0.05%) and blocked using PBS-5% no-fat milk. Human serum samples were thawed at room temperature, diluted (1:100 unless otherwise indicated), and incubated in blocking buffer for 1h (with vortexing) before plating. Serum samples were incubated overnight at 4°C before washing, as before. Secondary HRP-conjugated anti-human antibodies were diluted in blocking buffer and incubated with samples for 1 hour at room temperature. Plates were washed a final time before development with TMB Stabilized Chromogen (Invitrogen). The reaction was stopped using 1M sulphuric acid and optical density (OD) values were measured at 450 nm using an Asys Expert 96 ELISA reader (Biochrom Ltd.). Secondary antibodies (all from Southern Biotech) and dilutions used: goat anti-human IgG (2014-05) at 1:10,000; goat anti-human IgM (2020-05) at 1:1000; goat anti-human IgA (2050-05) at 1:6,000. All assays of the same antigen and isotype were developed for their fixed time and samples were randomized and run together on the same day when comparing binding between PCR+ individuals. Negative control samples were run alongside test samples in all assays and raw data were log transformed for statistical analyses.

### *In vitro* virus neutralisation assay

Pseudotyped viruses were generated by the co-transfection of HEK293T cells with plasmids encoding the SARS-CoV-2 spike protein harboring an 18 amino acid truncation of the cytoplasmic tail^14^; a plasmid encoding firefly luciferase; a lentiviral packaging plasmid (Addgene 8455) using Lipofectamine 3000 (Invitrogen). Media was changed 12-16 hours post-transfection and pseudotyped viruses harvested at 48- and 72-hours, filtered through a 0.45 µm filter and stored at −80°C until use. Pseudotyped neutralisation assays were adapted from protocols validated to characterize the neutralization of HIV, but with the use of HEK293T-*ACE2* cells. Briefly, pseudotyped viruses sufficient to generate ∼100,000 RLUs were incubated with serial dilutions of heat-inactivated serum for 60 min at 37°C. Approximately 15,000 HEK293T-ACE2 cells were then added to each well and the plates incubated at 37°C for 48 hours. Luminescence was measured using Bright-Glo (Promega) according to the manufacturer’s instructions on a GM-2000 luminometer (Promega) with an integration time of 0.3s. The limit of detection was at a 1:45 serum dilution.

### IL-6 cytometric bead array

Serum IL-6 levels were measured in a subset of PCR+ serum samples (*n=*64) using an enhanced sensitivity cytometric bead array against human IL-6 from BD Biosciences (Cat # 561512). Protocols were carried out according to the manufacturer’s recommendations and data acquired using a BD Celesta flow cytometer.

### Statistical analysis of SARS-CoV-2 PCR+ data

All univariate comparisons were performed using non-parametric analyses (Kruskal-Wallis, stratified Mann-Whitney, hypergeometric exact tests and Spearman rank correlation), as indicated, while multivariate comparisons were performed using linear regression of log transformed measures and Wald tests. For multivariate tests, all biochemical measures (IL-6, PSV ID50 neut., IgG, IgA, IgM) were log transformed to improve the symmetry of the distribution. As “days since first symptom” and “days since PCR+ test” are highly correlated, we cannot include both in any single analysis. Instead, we show results for one, then the other (Supp. Table 1).

### Probabilistic algorithms for classifying antibody positivity

Prior to analysis, each sample OD was standardized by dividing by the mean OD of “no sample controls” on that plate or other plates run on the same day. This resulted in more similar distributions for 2019 blood donor samples with 2020 blood donors and pregnant volunteers, as well as smaller coefficients of variation amongst PCR+ COVID patients for both SPIKE and RBD.

Our probabilistic learning approach consisted of evaluating different algorithms suited to ELISA data, which we compared through ten-fold cross validation (CV): logistic regression (LOG), linear discriminant analysis (LDA), support vector machines (SVM) with a linear kernel, and quadratic SVM (SVM2). Logistic regression and linear discriminant analysis both model log odds of a sample being case as a linear equation with a resulting linear decision boundary. The difference between the two methods is in how the coefficients for the linear models are estimated from the data. When applied to new data, the output of logistic regression and LDA is the probability of each new sample being a case. Support vector machines is an altogether different approach. We opted for a linear kernel, once again resulting in a linear boundary. SVM constructs a boundary that maximally separates the classes (i.e. the margin between the closest member of any class and the boundary is as wide as possible), hence points lying far away from their respective class boundaries do not play an important role in shaping it. SVM thus puts more weight on points closest to the class boundary, which in our case is far from being clear. Linear SVM has one tuning parameter *C*, a cost, with larger values resulting in narrower margins. We tuned *C* on a vector of values (0.001, 0.01, 0.5, 1, 2, 5, 10) via an internal 5-fold CV with 5 repeats (with the winning parameter used for the final model for the main CV iteration). We also note that the natural output of SVM are class labels rather than class probabilities, so the latter are obtained via the method of Platt^28^.

We considered three strategies for cross-validation: i) random: individuals were sampled into folds at random, ii) stratified: individuals were sampled into folds at random, subject to ensuring the balance of cases:controls remained fixed and iii) unbalanced: individuals were sampled into folds such that each fold was deliberately skewed to under or over-represent cases compared to the total sample. We sought a method with performance that was consistently good across all cross-validation sampling schemes, because the true proportion of cases in the test data is unknown, and we want a method that is not overly sensitive to the proportion of cases in the training data. We chose to assess performance using sensitivity and specificity, as well as consistency.

Given the good performance of all learners (described in the results), we considered the prediction surface associated with each SVM, LDA, SVM-LDA ensemble, and the standard 3-SD, 6-SD hard decision boundaries. Note that while methods trained on both proteins can draw decision contours at any angle, SD methods are limited to vertical or horizontal lines. We can see that success, or failure, of the SD cut-offs depends on how many positive and negative cases overlap for a given measure (S or RBD) in the training sample. In the training data the two classes are nearly linearly separable when each protein is considered on its own, which explains good performance of 3-SD and 6-SD thresholds. However, the test data contain many more points in the mid-range of S-RBD, which makes hard cut-offs a problematic choice for classifying test samples.

We trained the learners on all 733 training samples and used these to predict the probability of anti-SARS-CoV-2 antibodies in blood donors and pregnant volunteers sampled in 2020. We inferred the proportion of the sampled population with positive antibody status each week using multiple imputation. We repeatedly (1,000 times) imputed antibody status for each individual randomly according to the ensemble prediction, and then analyzed each of the 1,000 datasets in parallel, combining inference using Rubin’s rules, derived for the Wilson binomial proportion confidence interval^29^.

## Data Availability

Data generated as part of the study, along with custom code for statistical analyses, is openly available via our GitHub repositories: https://github.com/MurrellGroup/DiscriminativeSeroprevalence/ and https://github.com/chr1swallace/seroprevalence-paper.

## Data and code availability statement

Data generated as part of the study, along with custom code for statistical analyses, is openly available via our GitHub repository: https://github.com/chr1swallace/elisa-paper.

## Author contributions

GKH and XCD designed the study and wrote the manuscript with input from co-authors. JA, TA, JD, SM, GB, MA and SA provided the study serum samples and clinical information. LH, LPV, AMM, DJS, KCI, BM and GM generated SARS-CoV-2 antigens and pseudotyped viruses. MF and XCD developed the ELISA protocols and XCD generated the data. DJS and BM performed the neutralization assay. CW and NFG executed machine learning approaches and statistical analyses, with input from MCh and BM. MA, SK, PP, MM, JC, MCo and JR carried out wet lab experiments and assisted with data analysis.

## Acknowledgments

We would like to thank the study participants and attending clinical teams. Secondly, we extend our thanks to Björn Reinius, Marc Panas, Julian Stark, Remy M. Muts and Darío Solis Sayago for their input and discussion. Funding for this work was provided by a Distinguished Professor grant from the Swedish Research Council (agreement 2017-00968) and NIH (agreement 400 SUM1A44462-02). CW and NFG are funded by the Wellcome Trust (WT107881) and MRC (MC_UP_1302/5). For the purpose of Open Access, the author has applied a CC BY public copyright licence to any Author Accepted Manuscript version arising from this submission.

---

**Figure S1:**
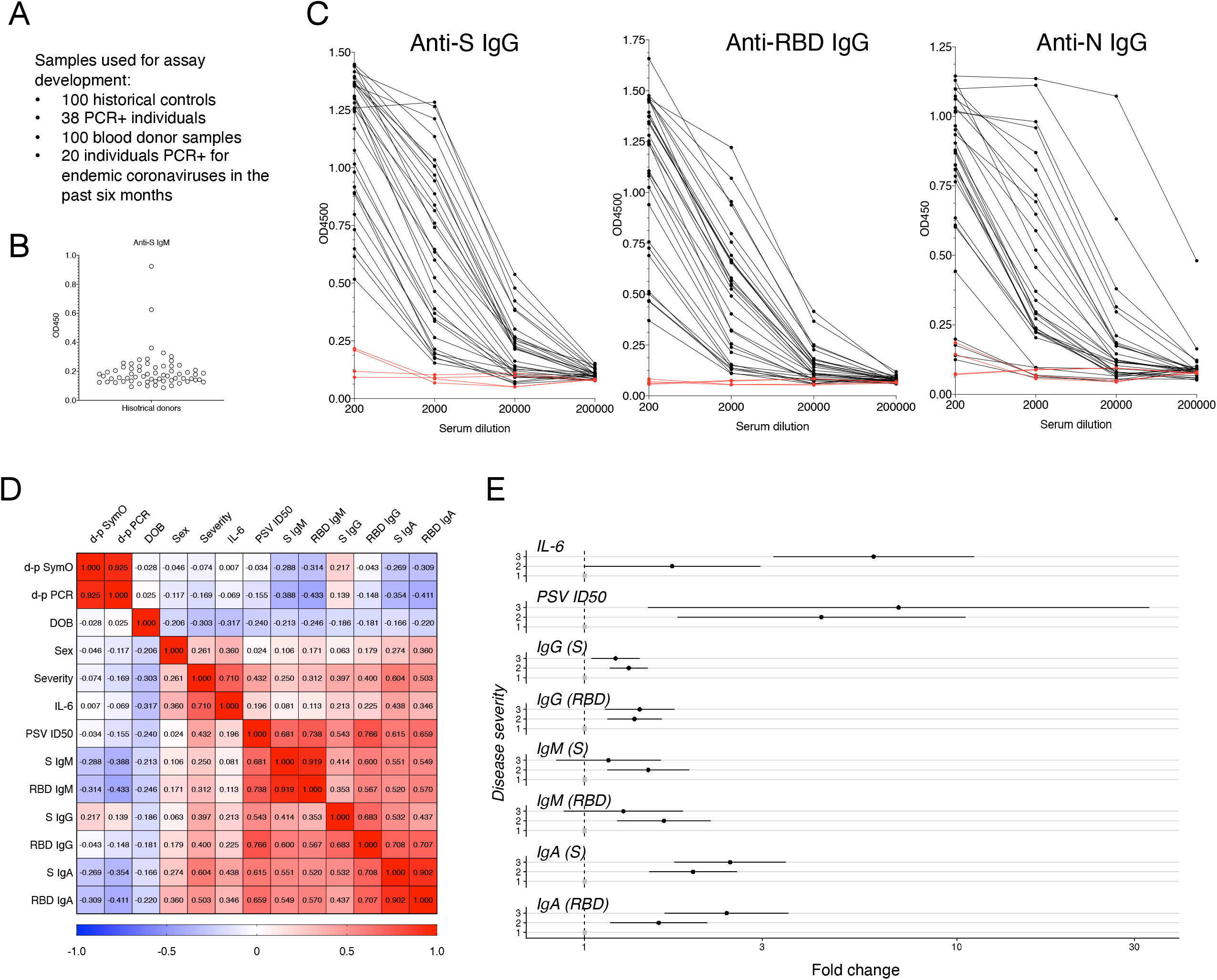
Antibody phenotypes in PCR+ individuals. **(A)** Study samples used for assay development. **(B)** Anti-S IgM reactivity observed in a random subset of historical controls. Binding was confirmed in these samples in an independent experiment. No reproducible IgG reactivity to S trimers of the RBD was observed across all historical controls in the study. **(C)** Serial dilution of *n=*30 random PCR+ individuals. ECV+ (*n=*4) controls are shown in red. **(D)** Spearman’s rank correlation of PCR+ dataset features and antibody levels. DOB - *date of birth*; d-p SymO - *days post-symptom onset*; d-p PCR – *days post SARS-CoV-2+ PCR*; PSV ID50 – *neutralizing titer*. **(E)** Adjusted fold-change compared to Category 1 PCR+ individuals. The effects of age (DOB), sex, days from PCR test were considered.

**Figure S2:**
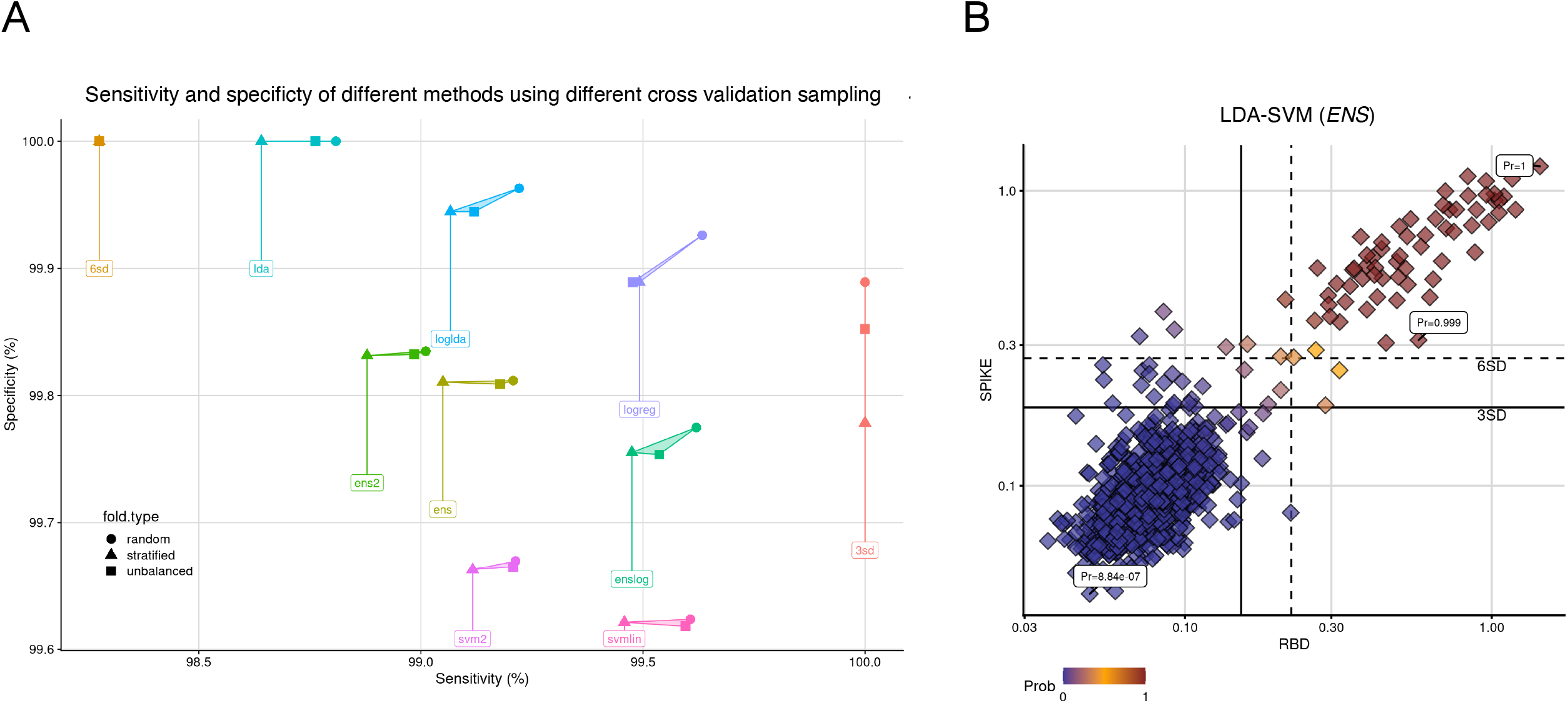
Performance of different probabilistic approaches. **(A)** Comparisons of specificity and sensitivity for the different probabilistic methods (and 3 and 6 SD thresholding) using different cross-validation strategies. **(B)** *ENS* probabilities when applied to healthy donor test data, providing a highly sensitive, specific and consistent multi-dimensional solution to the problem of low responders, and assigning each data point a probability of being positive.

